# Image and structured data analysis for prognostication of health outcomes in patients presenting to the Emergency Department during the COVID-19 pandemic

**DOI:** 10.1101/2021.07.07.21260097

**Authors:** Liam Butler, Ibrahim Karabayir, Mohammad Samie Tootooni, Majid Afshar, Ari Goldberg, Oguz Akbilgic

## Abstract

**Background:** Patients admitted to the emergency department (ED) with COVID-19 symptoms are routinely required to have chest radiographs and computed tomography (CT) scans. COVID-19 infection has been directly related to development of acute respiratory distress syndrome (ARDS) and severe infections lead to admission to intensive care and can also lead to death. The use of clinical data in machine learning models available at time of admission to ED can be used to assess possible risk of ARDS, need for intensive care unit (ICU) admission as well as risk of mortality. In addition, chest radiographs can be inputted into a deep learning model to further assess these risks.

**Purpose:** This research aimed to develop machine and deep learning models using both structured clinical data and image data from the electronic health record (EHR) to adverse outcomes following ED admission.

**Materials and Methods:** Light Gradient Boosting Machines (LightGBM) was used as the main machine learning algorithm using all clinical data including 42 variables. Compact models were also developed using 15 the most important variables to increase applicability of the models in clinical settings. To predict risk of the aforementioned health outcome events, transfer learning from the CheXNet model was implemented on our data as well. This research utilized clinical data and chest radiographs of 3571 patients 18 years and older admitted to the emergency department between 9^th^ March 2020 and 29^th^ October 2020 at Loyola University Medical Center.

**Main Findings:** Our research results show that we can detect COVID-19 infection (AUC = 0.790 (0.746-0.835)) and predict the risk of developing ARDS (AUC = 0.781 (0.690-0.872), ICU admission (AUC = 0.675 (0.620-0.713)), and mortality (AUC = 0.759 (0.678-0.840)) at moderate accuracy from both chest X-ray images and clinical data.

**Principal Conclusions:** The results can help in clinical decision making, especially when addressing ARDS and mortality, during the assessment of patients admitted to the ED with or without COVID-19 symptoms.

## 1. Introduction

The novel SARS-CoV-2 (COVID-19) virus has quickly spread globally and was classified as a world pandemic by the World Health Organization (WHO) on 11^th^ of March 2020 (Cucinotta & Vanelli, 2020). There are currently more than 2 million people infected with this virus. It is reported that this virus has high probability of causing severe acute respiratory syndrome (ARDS); therefore, requires early identification and treatment(Cucinotta & Vanelli, 2020). Since the outbreak of the COVID-19 pandemic, almost 124 million people have been infected and more than 2.73 million people have died from COVID-19 infection (WHO, 2021). A substantial number of infected individuals arrive at the emergency department (ED) with hypoxic respiratory failure from COVID-19, with greater prevalence among individuals 65 years of age and older (Bhatnagar et al., 2020; Chavez et al., 2020).

Testing for COVID-19 virus has evolved with multiple assays achieving high sensitivity and greater distribution of testing, including general symptom assessment and saliva. Part of routine care for patients presenting to the emergency department with COVID-19 symptoms include chest imaging with radiographs and computed tomography (CT) scans. Image-based deep learning models can rapidly identify outcomes of the infection in different organs such as lungs, which can be substantial aids to clinicians for time-sensitive diagnostics (Castiglioni et al., 2020). COVID-19 infections have been linked to the development of acute respiratory distress syndrome (ARDS), which results in decreased lung space and loss of lung tissue aeration (Ranieri et al., 2012). The use of chest radiographs can be inputted into a deep learning model (Gozes et al., 2020) to predict the probability of infection in each patient as well as facilitate triage to the intensive care unit (ICU) and also signal the risk for mortality.

This research aimed to develop machine and deep learning models using both structured clinical data and image data from the electronic health record (EHR) to discriminate the following events: (1) COVID-19 infections, (2) acute respiratory distress syndrome in patients with and without COVID-19; (3) need for ICU admission; (4) and in-hospital death. Further deep learning models to determine probability of (2) to (4) were also developed using image analysis from chest radiographs. An additional aim of this research was to develop compact models, with similar accuracy to the ‘full’ models, using a smaller number of variables to increase generalizability of model implementation in clinical settings.

## 2. Materials & Methods

### 2.1 Cohort

The cohort used in this study included patients admitted to Loyola University Medical Centre, Maywood, Illinois, USA who were admitted to emergency department between 9^th^ March 2020 and 29^th^ October 2020. The total number of patients included in the study was 3571. Inclusion criteria was that all adults (>18yo) encounters in the emergency department who were tested for COVID-19 and received chest imaging. This research was approved by an Institutional Review Board (IRB).

### 2.2 Outcomes

The main outcome of this research was the risk prediction of i) COVID-19 infection, ii) acute respiratory distress syndome (ARDS), iii) requirement for ICU admission and iv) risk of mortality at time of emergency department (ED) admission. COVID-19 infection was identified from laboratory tests. ARDS, following the Berlin definition, is an acute diffuse lung injury that can increase pulmonary vascular permeability. ARDS can be identified through chest radiographs due to the development of hypxemia and bilateral radiographic opacities (Azzam et al., 2009; Koenig et al., 2011; Ranieri et al., 2012). The algorithm included vital signs, laboratory data, ventilator data, and keywords from the chest imaging that met the requirement for hypoxemia and bilateral infiltrates without a primary cardiogenic etiology for edema. The algorithm was updated from the original studies to include CT reports, minimum peak end expiratory pressure of 5cm H2O on the ventilator and meeting the criteria within seven days of hospital presentation for primary ARDS.

### 2.3 Risk Factors

A total of 42 structured data variables from the EHR were included as risk factors in the first machine learning models. The cohort data included demographics such as age, gender, race and ethnicity, and initial measurement of oxygen saturation levels. The data also included comorbidities that was present on admission, including but not limited to heart disease (cardiovascular disease, congestive heart failure etc.), lung disease (chronic obstructive pulmonary disease (COPD), pulmonary circulation disorders etc.), liver and renal diseases as well as abusive drug or alcohol intake. This data was obtained using pre-specified ICD10 codes. A summary statistics of the included variables are provided in Table S.1 in Supplementary Materials. Our dataset was comprehensive and only had minimal amounts of missing data with 30 patients having no recorded ethnicity and 2 patients having no recorded first oxygen levels. Due to the low amount of missing data, these two variables were imputed using the most frequent category or the median of the cohort.

### 2.4 Chest Imaging Data

This research also used chest radiographs, imported in DICOM format. One frontal radiograph for each patientwas exported in .*jpeg* format. The dataset included 3571 images, with 1 chest radiograph per patient. Pre-trained deep learning models on X-ray and CT scans already exist (e.g. CheXNet) (Rajpurkar et al., 2017) and can be used as a baseline to assess risk associated with ARDS, need for ICU admission and possibly risk of mortality following COVID-19 infection (American College of Chest Physicians, 2020). The chest radiographs were rescaled to the same size (between 0 and 1) implemented in the original CheXNet model (Rajpurkar et al., 2017). The images were resized to 224 × 224 size for training the transfer learned fine-tuned CheXNet-DenseNet121 model. An image data generator was used to load the images and the associated labelling, stored in a .*csv* file, for each of the 4 outcomes. Image data was augmented during training to ensure that the model can classify images appropriately in cases when images either contain noise or are shifted or rotated as well as to reduce overfitting of the data.

### 2.5 Outcome Prediction using Clinical Risk Factors

We randomly split the study cohort into 80% for model building and 20% of the data retained as a holdout test set. Hyperparamaters for a Light Gradient Boosting Machines (LGBM) were tuned using 5-fold cross validation on the 80% training set to achieve the optimal area under the receiver operating characteristics (AUC). The integration of cross-validation methods have been widely used in machine learning studies and have shown that this strategy can increase model performance when compared to a 80%/20% training-holdout split with no cross-validation (Yadav & Shukla, 2016). The trained model was tested on the 20% holdout data. All the models comparisons and evalutions were based on AUC statistics obtained on the holdout data. Models were compared using DeLong Test, a non-paramateric test for comparison of independent AUCs of models (DeLong et al., 1988). For each outcome, a ‘shapley additive explanations’ (SHAP) variable importance analysis (Molnar, 2020) was performed on each model and the 15 top most important variables identified as best predictors for each of the 4 outcomes. SHAP is a method to explain individual predictions as well as show the global positive and negative relationships of the predictors with the outcome (Molnar, 2020). With SHAP, global interpretations of the model are consistent with the local explanations (for each observation), since the importance analysis is based on the combined ‘Shapley’ values of the global interpretations (Molnar, 2020). In this research we provided SHAP summary plots which combine the global importance of the variables and the effects of these variable with the outcome.

### 2.6 Outcome Prediction from radiograph images using CheXNet

CheXNet is a pretrained convolutional neural network (CNN) model to process chest radiographs (X-Ray) images to detect 14 classes namely: Atelectasis, Cardiomegaly, Effusion, Infiltration, Mass, Nodule, Pneumonia, Pneumothorax, Consolidation, Edema, Emphysema, Fibrosis, Pleural Thickening and Hernia (Rajpurkar et al., 2017). CheXNet model was developed on 120,000 chest radiograph images available at “https://github.com/jrzech/reproduce-chexnet” and “https://github.com/brucechou1983/CheXNet-Keras“, with the full image data set available from the open access National Institute of health (NIH) database at https://nihcc.app.box.com/v/ChestXray-NIHCC (Wang et al., 2017). We used existing CheXNet model on our chest radiograph images and obtained risk predictions for all 14 categories. We then used these 14 predictions as individual predictors of our four outcomes to identify possible CheXNet categories associated with our outcomes of interest. The highest resulting AUCs from each of the 14 classes were identified and used. This means that we further developed models that also included a select number of predicted lung disease classes, namely consolidation, infiltration and pneumonia, adopted from CheXNet predictions that are associated with higher risks, rather than using all 14 classes from CheXNet predictions. The model used pre-tarined weights from ImageNet and training utilized DenseNet121 architecture, with an initial learning rate of 0.001, reducing with a factor of 10 when validation loss did not decrease after each epoch. The imaging training model used a batch size of 64.

### 2.7 Transfer Learning to create CheXNet-Cov19

Our study then used transfer learning to obtain a CheXNet based deep learning model, namely CheXNet-Cov19, that predicts risk for our 4 outcomes of interest (COVID-19 infection, ARDS, ICU admission and risk of mortality), with each outcome being a probability between 0 and 1 for the associated risk. To do that, we first altered the CheXNet arthitecture by replacing the 14 nodes outcome layer with four nodes representing COVID-19 infection, ARDS development, ICU admission and mortality. We then initialized all parameters of this new artitecture from CheXNet except the last fully connected layer. We then re-trained and fine-tuned this novel CNN model on 70% of our radiograph images while using 10% of images for validation. The weights for each class for each outcome was based on total counts and class positive counts. The initial learning rate was set to 0.0001, reducing with a factor of 10 when validation loss did not decrease after each epochs. The batch size was set to 64. The final trained model was tested on the same 20% holdout dataset that was used in earlier predictive models.

### 2.8 Integration of Chest Radiographs and Clinical Data

We then integrated radiograph-based predicted risks to each of the final models built on clinical data. We did this by investigating combinations (or ensembles) of various risk predictions and/or risk factors using the machine learning algorithm Light Gradient Boosting Machines (LGBM). In this ensemble approach, we built final prediction models on the same 80% model building dataset (previously discussed) and evaluated the models on the same 20% holdout dataset for streamlined comparisons.

The models and related analyses were performed using the Python programming language and the associated code will be available in a github repository.

## 3. Results

The cohort was composed of 3,571 patient ecnounters, of which 1,907 (53.40%) were male, 1,605 (44.95%) were of white race, and 944 (26.44%) of Hispanic ethnicity. The mean age of the cohort, with standard deviation, was 56.2S3±20.54. Oxygen levels were taken when patients first arrived at ED with mean oxygen saturation of 96.27%±5.35. There were 789 patients (22.09%) diagnosed with COVID-19 infection with laboratory confirmation through assays, 260 patients (7.28%) developed ARDS and 963 patients (26.97%) were admitted to an ICU. From those admitted to an ICU, 435 patients were admitted directly to ICU with the remaining 528 patients being admitted to another ward first. In-hospital dearh occurred in 293 patients (8.20%), with 212 patients dying in the ICU. In addition, from the 963 patients admitted to ICU, 245 had developed ARDS. The summarized statistics are detailed in Table 1 seperately for COVID-19 positive and negative patients. The details of all 42 risk factors are provided in Supplementary Materials Table 1.

**Table 1.** Summary statistics comparing COVID-19 positive and COVID-19 negative patients who i) developed ARDS, ii) were admitted to ICU and iii) died.

|  | Total<br>(N = 3571) | COVID positive<br>(N = 789) | COVID negative<br>(N = 2782) | p-value |
| --- | --- | --- | --- | --- |
| No. patients developed ARDS, N (%) | 260 (7.28) | 101(12.80) | 159 (5.72) | <0.001 |
| No. patients admitted to ICU, N (%) | 963 (26.97) | 243 (30.80) | 720 (25.88) | 0.006 |
| No patients who died, N (%) | 293 (8.20) | 91(11.53) | 202 (7.26) | <0.001 |

### 3.1 Outcome Prediction using Clinical Risk Factors

Models built on the full clinical data set (all 42 risk factors) resulted in moderate accuracies to predict COVID-19 infection with an AUC of 0.790 (0.746-0.835), ARDS with an AUC of 0.753 (0.675-0.831), ICU transfer with an AUC of 0.675 (0.620-0.713), and in-hospital death with an AUC of 0.683 (0.606-0.761). A set of 15 top most important predictors for all four outcomes from SHAP variable importance analysis is provided in Table S.2 as supplementary material. We further re-built compact models using only those top 15 clinical risk factors seperatley for each outcome of interest. The use of more compact models can allow for generalisability in situations where medical information is not as comprehensive in other clinical settings. The compact models provided prediction accuracies with an AUC of 0.775 (0.730-0.821) for COVID-19 infection, 0.721 (0.641-0.802) for ARDS, 0.658 (0.611-0.702) for ICU admission and AUC of 0.755 (0.669-0.841) for mortality. Compared to models using all 42 variables, DeLong test showed that the compact models for need of ICU admission and mortality were significantly better (p<0.05) but not significantly different for COVID-19 infection (p=0.762) and ARDS (p=0.071). Figures 1a-d provide the variable importance analysis results for models aimed to predict risk for COVID-19 infection, ARDS, need for ICU admission and risk of mortality based on the 15 most important clinical predictors. First oxygen (O_2_) levels are the most important predictors for COVID-19 infection (Figure 1a), ARDS (Figure 1b) and ICU admission (Figure 1c), and second-most important predictor for risk of mortality (Figure 1d) superceded by age. Interestingly, both race and Hispanic ethnicity are amongst the most important contributors to the COVID-19 infection models. COVID-19 infection is a major contributor, as expected, to the development of ARDS but is less important in ICU admission and risk of mortality. As expected, age is an important predictor for each of the four outcomes, albeit with different levels of importance.

**Figure 1a.**
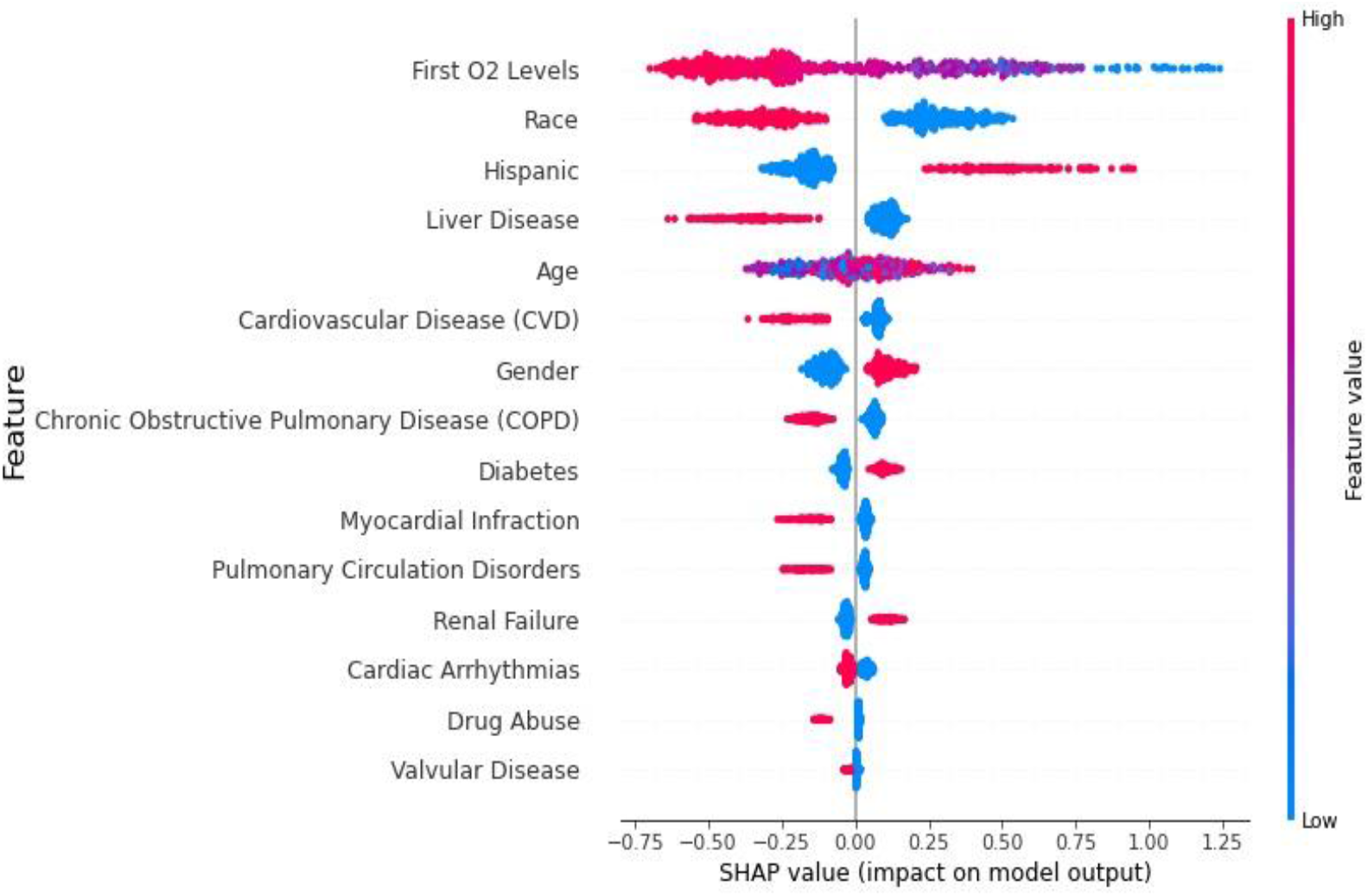
SHAP Variable importance analysis using 15 clinical variables to predict risk of COVID-19 infection.

**Figure 1b.**
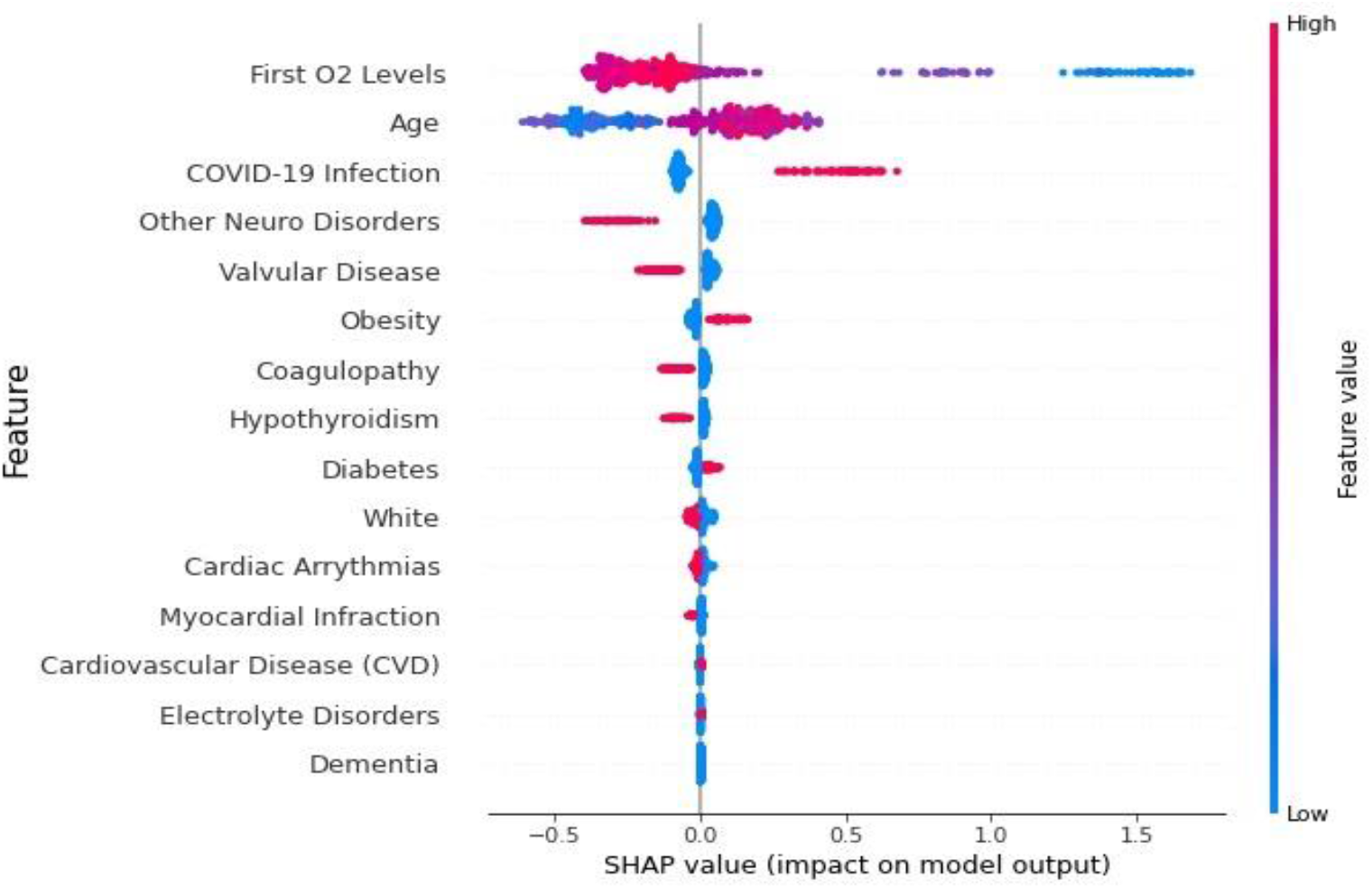
SHAP Variable importance analysis using 15 clinical variables to predict risk of ARDS.

**Figure 1c.**
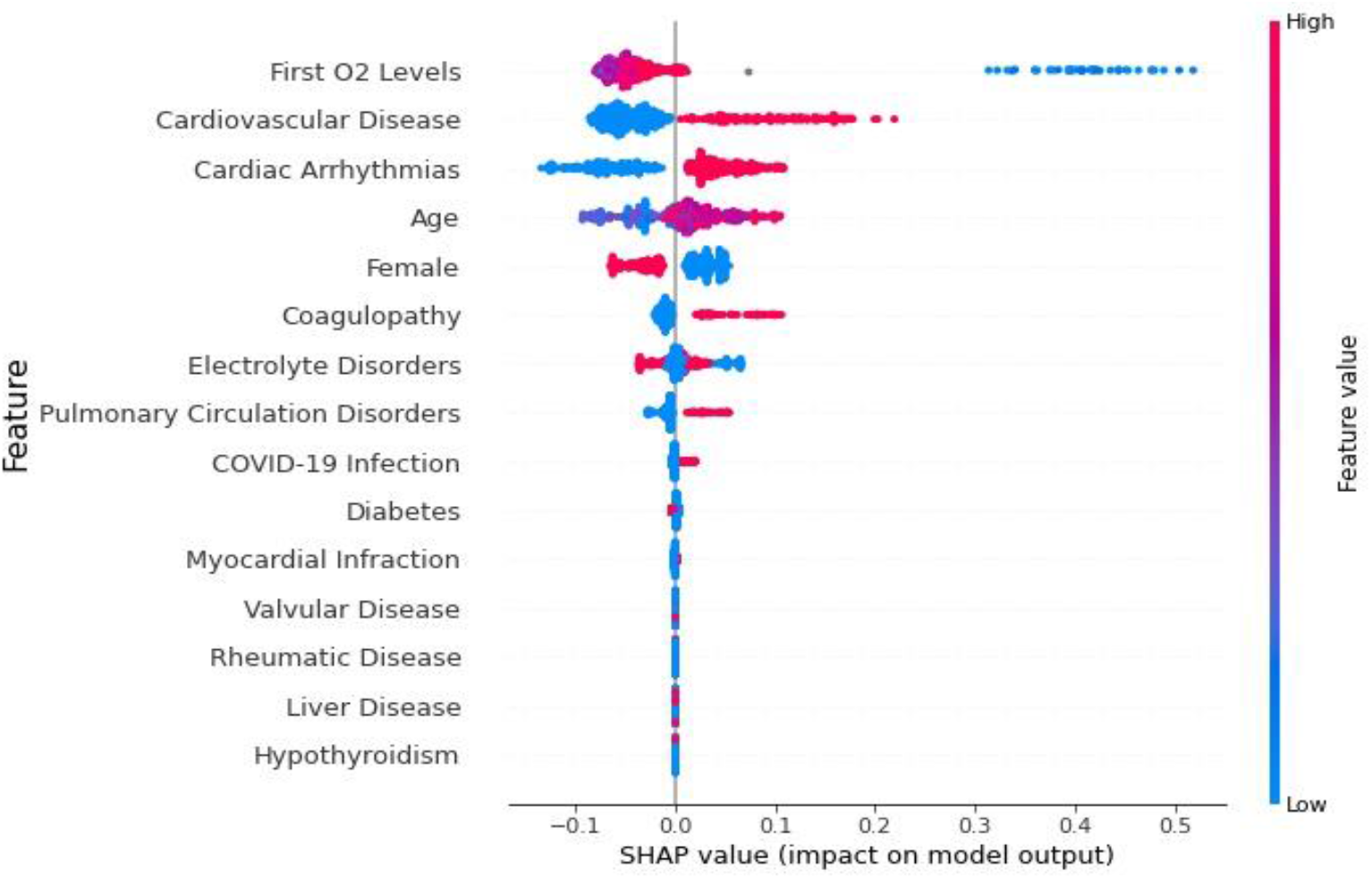
SHAP Variable importance analysis using 15 clinical variables to predict risk or need of ICU admission.

**Figure 1d.**
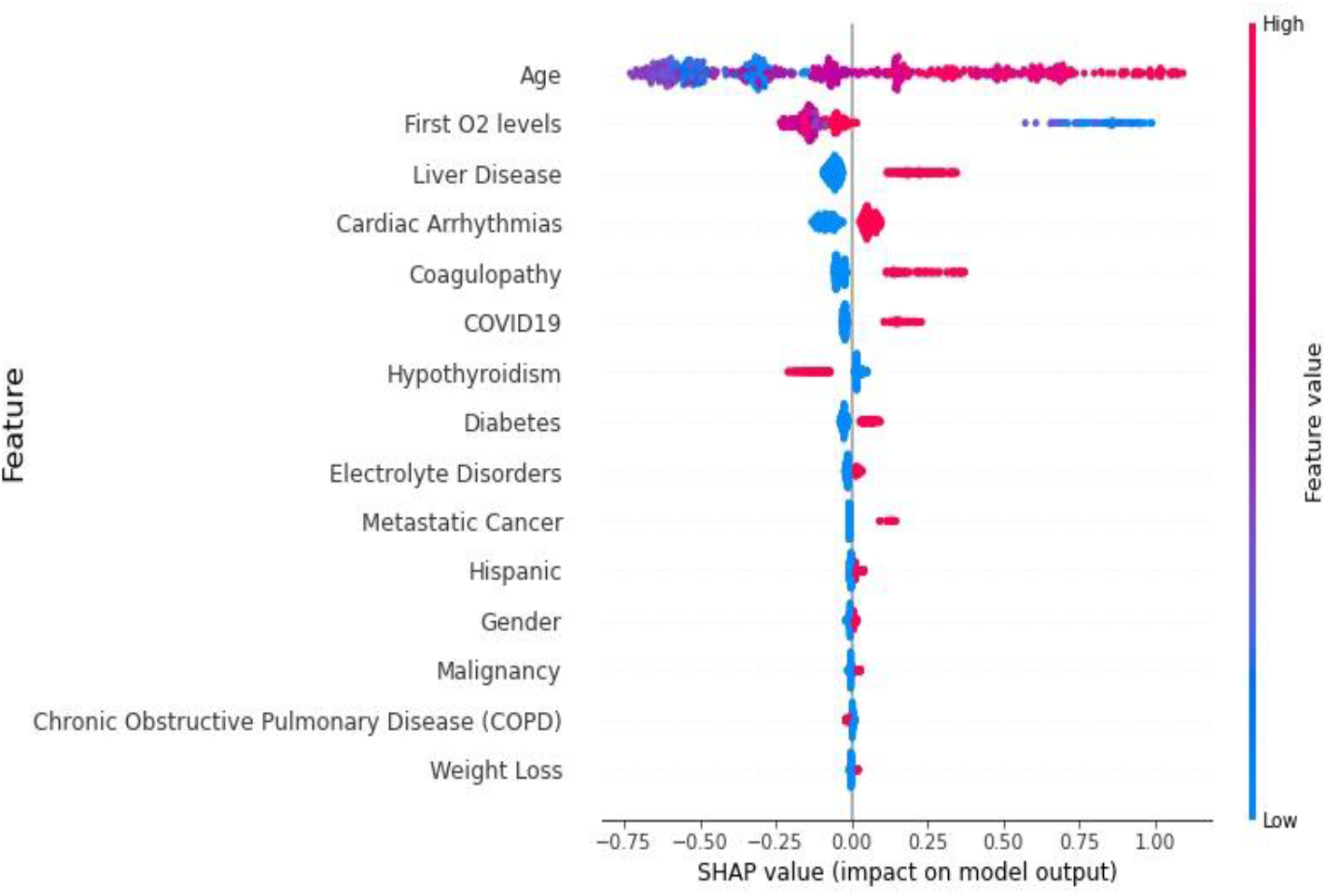
SHAP Variable importance analysis using 15 clinical variables to predict risk of mortality.

### 3.2 Outcome Prediction from X-Ray images using CheXNet

The AUCs to predict ARDS were 0.624 for Atelectasis, 0.547 for Cardiomegaly, 0.625 for Effusion, 0.735 for Infiltration, 0.557 for Mass, 0.540 for Nodule, 0.702 for Pneumonia, 0.696 for Pneumothorax, 0.711 for Consolidation, 0.697 for Edema, 0.499 for Emphysema, 0.536 for Fibrosis, 0.586 for Pleural Thickening and 0.471 for Hernia. The categories with highest AUC for ARDS prediction, namely infiltration, pneumonia and consolidation were used in further models. This was deemed fit since all three classes also share similarities in disease with ARDS. While this type of data will not be available at time of admission to ED, the implementation of CheXNet and image-based models can be available at the time when the chest radiograph is available.

We built additional models to predict each of the 4 outcomes in our aims by adding the predictions for three named CheXNet categories. The inclusion of predicted infiltration, pneumonia and consolidation to the defined 15 variables resulted in prediction accuracies with an AUC of 0.748 (0.681-0.814) for COVID-19 infection, 0.694 (0.596-0.793) for ARDS, 0.675 (0.611-0.702) for ICU transfer and 0.748 (0.661-0.835) for mortality. Compared to the compact models using only 15 clinical variables, using DeLong Test, the AUCs were significanly higher for all of the outcomes under study with p<0.001.

We also built respective models using demographic data, first oxygen levels, and covid infection (for ARDS, ICU admission and mortality) together with infiltration, pneumonia and consolidation to assess the power of these X-Ray specific predictors on predicting each of the 4 classes. The use of, solely, demograpics, first oxygen levels and the three CheXNet predicted classes resulted in AUC of 0.730 (0.663-0.797). When COVID-19 infection was added as a predictor for ARDS, ICU admission and Mortality, AUC scores for ARDS increased to 0.702 (0.604-0.800) and 0.750 (0.664-0.837) for risk of mortality. The results for each developed model are provided in Table 2. Furthermore, calibration was performed for the best machine learning models using clinical data, namely COVID-19 infection, ARDS and ICU using all 42 variables and using 15 clinical variables for risk of mortality. Figures S.1a-d in Supplementary material show the calibration curves. Calibration of the models only minimally improved, if at all, predictions from the associated models. We acknowledge that in some cases the presented models can over- or under-estimate the predicted probabilities for each outcome using clinical variables (e.g. FigureS.1b and d). This could be due to the imbalance between positive cases and the rest of the patient cohort within the dataset for each of the four outcomes in the study.

**Table 2.** Models to predict covid infection, ARDS, ICU admission and risk of mortality using i) all available clinical variables, ii) 15 top clinical variables, iii) 15 top clinical variables in addition to the three predicted classes from CheXNet and iv) demographics + first oxygen levels + 3 predict CheXNet classes.

| **Model Type** | **COVID-19** | **Acute Respiratory Distress Syndrome (ARDS)\*** | **ICU Admission\*** | **Mortality\*** |
| --- | --- | --- | --- | --- |
| All 42 clinical variables | 0.790 (0.746-0.835) | 0.753 (0.675-0.831) | 0.675 (0.620-0.713) | 0.683 (0.606-0.761) |
| Top 15 clinical variables | 0.775 (0.730-0.821) | 0.721 (0.641-0.802) | 0.658 (0.611-0.702) | 0.755 (0.669-0.841) |
| 15 clinical variables + CheXNet risk predictions of (consolidation + infiltration + pneumonia) | 0.748 (0.681-0.814) | 0.694 (0.596-0.793) | 0.675 (0.622-0.727) | 0.748 (0.661-0.835) |
| Demographics + first oxygen levels + CheXNet risk predictions of (consolidation + infiltration + pneumonia) | 0.730 (0.663-0.797) | 0.702 (0.604-0.800) | 0.657 (0.603-0.710) | 0.750 (0.664-0.837) |
| *Models include COVID-19 infection binary status as a predictor |  |  |  |  |

### 3.3 Transfer Learning to create CheXNet-Cov19

CheXNet-Cov19 model provided prediction accuracies with an AUC of 0.712 (0.627-0.797) for COVID-19 infection, 0.741 (0.658-0.824) for risk of ARDS, 0.665 (0.578-0.752) for ICU admission and 0.759 (0.678-0.840) for mortality (Table 3). Table 3 also highlights and compares the results obtained when building different models on i) all variables + predicted risks, ii) 15 top variables + predicted risks and iii) 15 top variables + predicted risks + predicted infiltration, consolidation and pneumonia.

**Table 3.** Models to predict ARDS, ICU admission and risk of mortality using i) transfer learning from CheXNet, ii) all clinical variables + CheXNet risk predictions, iii) 15 top clinical variables + the predicted risk and iv) top 15 clinical variables in addition to predicted risks as well as risk of consolidation, infiltration and pneumonia (from initial CheXNet model)

| Model Type | COVID-19 | Acute Respiratory Distress Syndrome (ARDS) | ICU Admission | Mortality |
| --- | --- | --- | --- | --- |
| CheXNet-Cov19 using chest X-Rays only | 0.712 (0.627-0.797) | 0.741 (0.658-0.824) | 0.665 (0.578-0.752) | 0.759 (0.678-0.840) |
| CheXNet-Cov19 Predicted risk* + All clinical variables |  | 0.776 (0.685-0.867) | 0.663 (0.609–0.716) | 0.736 (0.691-0.781) |
| CheXNet-Cov19 Predicted Risk + 15 clinical variables * |  | 0.781 (0.690-0.872) | 0.647 (0.593-0.701) | 0.751 (0.664-0.837) |
| CheXNet-Cov19 predicted Risk + 15 clinical variables + consolidation + infiltration + pneumonia * |  | 0.728 (0.632-0.824) | 0.675 (0.622-0.727) | 0.758 (0.672-0.844) |
| *Models include COVID-19 infection status |  |  |  |  |

The inclusion of predicted risk to the model built on all the predictors resulted in AUC of 0.776 (0.685-0.867) for ARDS, AUC of 0.663 (0.609– 0.716) for need of ICU admission and AUC of 0.736 (0.69-0.781) for risk of mortality. Similar results were obtained when the number of variables within the model was 15 variables + predicted risks. The inclusion of risk of infiltration, consolidation and pneumonia predicted from the CheXNet CNN resulted in AUCs of 0.728 (0.632-0.824), 0.675 (0.622-0.727) and 0.758 (0.672-0.844) for ARDS, need of ICU admission and mortality respectively.

## 4. Discussion

The novel COVID-19 virus has taken the world by surprise and has been recognized as a pandemic due to the it’s ability of quick infection, transmission as well as mutation into more transmissible variants (Darby & Hiscox, 2021). Numerous scientific communities have taken to study the virus and its effects on human health with results showing that the virus predominantly attacks the lungs and was the cause of a large influx of patients to intensive care and a consequently very high mortality rate (Baud et al., 2020). Even more so, lung infections caused by this viral infection differs from ‘typical’ pneumonia, and identification of viral pneumonia has been reportedly difficult (Gibson et al., 2020). While testing for COVID-19 infections has progressed substantially to very rapid testing with the main aim of containing viral infection by encouraging people to self-quarantine, there has still be a need for early prediction of possible severe outcomes which can result in ICU admission and/or mortality if left undetected or untreated. People with severe infections and symptoms, especially those of older age, are generally admitted to the emergency department during which a series of chest X-rays are taken to be assigned appropriate treatment by a consulting physician if there was determinable acute respiratory distress syndrome (Dreher et al., 2020). However, there has been a recorded bottleneck between time of taking chest X-rays, diagnosis of severity of infection and admission to ICU, unless severe cases have been recognized by ambulatory services (Li et al., 2020).

In our study, we developed a series of models that used both clinical variables using machine learning as well as chest X-rays (in CNN) to try and predict risk of ARDS, the possible need for ICU admission, and risk of mortality. Our models were built on patient data available during admission to the emergency department as well as the utilization of transfer learning, using CheXNet (Rajpurkar et al., 2017), to predict lung-specific diseases most notably infiltration, consolidation and pneumonia. These 3 conditions were added to the most important risk predictors to assess risk of ARDS, need for ICU admission and risk of mortality.

Prediction of risk of ARDS resulted in a moderate AUC of 0.721 (0.641-0.802) using solely 15 short-listed clinical variables without the addition of predicted risk of infiltration, consolidation or pneumonia while risk of mortality was predicted with an AUC of 0.755 (0.669-0.841) without the addition of these risk variables. Variable importance analyses for each model shows that first oxygen levels and age are amongst the most important predictors. This is what was expected since COVID-19 infection reduces oxygen levels within the blood, as well as older patients being more susceptible to infection because of higher proportion of comorbidities (Niu et al., 2020). However, variable importance analysis for ARDS prediction models show that COVID-19 infection is indeed within the top three important predictors, conferring the association between COVID-19 viral infection and acute respiratory distress syndome, adding face validity to the models developed. On the other hand, variable importance analysis for models built to assess the need for ICU admission (Figure 1b), the first oxygen levels retain their importance followed by heart disease. When predicting risk of mortality, Figure 1c shows that Age is, unsurpringly, the largest contributor to mortality, followed by oxygen levels and liver and cardiac diseases. COVID-19 infections might have potentially had a role in increased risk of mortaltiy due to its higher importance shown in Figure 1c.

When these risk variables for infiltration, consolidation, or pneumonia were added to the models, the AUC did not change much for prediction of COVID-19 infection, while AUC for prediction of ARDS with inclusion of these external variables decreased to 0.694 (0.596-0.793), presumably due to the similarities and overlap between these three variables and the broad definition of acute respiratory distress syndrome (Weinacker & Vaszar, 2001). The addition of predicted risks for ARDS, ICU admission and mortality were added to the short-listed clinical variables in the initial models and results show a notable increase in accuracy for prediction of ARDS (0.781) while retained similarities to models built on solely short-listed clinical variables for mortality (0.751). However, we note that predicted risk of mortality solely from chest radiographs is slightly higher (0.759). The models built to predict possibility of mortality also include within them predicted risk of ARDS and need for ICU admission. The reason for this is because our developed models can be utilized at time of deployment of chest radiographs at first time of availability in a clinical setting, potentially helping clinicians determine best course of treatment. We acknowledge that admission to intensive care is somewhat subjective by the trained physician, but it would be possible to include the predicted risk in the model for increased precaution by decreasing the time spent at emergency department when the patient has a high noted risk of requiring intensive care (Ma & Vervoort, 2020).

From the implementation perspective, it is important to note that intensive care units during a pandemic can be overwhelmed. For future prospective analysis and potential implementation, we suggest using the compacted models that were developed to increase generalizability of model implementation and to reduce clinical burden, especially in clinical settings. The results from DeLong test showed that compact models with predicted risk transferred from image analysis are preferred over the original models and can better increase awareness and help clinicians in decision making to assess need for ICU admission and risk of mortality. There is additional scope for the utilization of incorporation of image analysis and prediction on mobile technologies such as smartphones. The use of predictive models incorporated into such technologies can potentially help clinicians to incorporate machine learning/deep learning efforts into the workflow.

## 5. Limitations

Our research has some limitations. First, it is a single site study, therefore, require an external validation before its use in clinical practice. Also, there is a data imbalance in the number of patients with COVID-19 infections and ARDS, which could have reduced the models’ training and performance when using clinical variables and image analysis. The data also, whilst a small proportion, included patients that had developed ARDS but did not have COVID-19 infection. In addition, prediction of ICU admission scored a comparatively lower AUC which could be because of ICU admission not directly related to the predictive variables that were collected and used. In addition, ICU admission is largely subjective based on clinicians’ professional advice in terms of severity of illness bed availability, hospitalization costs as well as varying substantially between hospitals (Robert et al., 2015).

## Conclusions

Our research results show that we can predict risk of COVID-19 infection, ARDS and mortality at moderate accuracy from both chest X-ray images as well as with the addition of clinical variables. This research offers clinical applicability in that a developed tool that can assess chest X-rays at time that they are taken and can substantially help clinicians in decision making as well as reduce burden on intensive care units. Since our dataset also included COVID-19 negative patients, it proposes a methodological approach of assessing ARDS and risk of mortality irrespective of COVID-19 status. Additionally, the use of solely X-ray data can potentially be improved with increased computational power, high batch size analysis as well as a larger cohort size.

## Data Availability

The models and related analyses were performed using the Python programming language and the associated code will be available in a Github repository.

## Acknowledgements

We would like to thank Loyola University Chicago, Loyola University Medical Centre and all associated personnel in their contirbution to data acquisiton.

## Supplementary Material

**Table S. 1.** Summary of the 42 clinical characteristics used in this study, as well as relationship with COVID-19 infections, ARDS, need for ICU admission and mortality.

|  | Variable | N (%) |
| --- | --- | --- |
| 1 | Age, mean±s.d. | 56.23±20.54 |
| 2 | AIDS-HIV | 21 (00.59) |
| 3 | Alcohol Abuse | 86 (2.41) |
| 4 | Blood Loss Anemia | 162 (4.54) |
| 5 | Cardiac Arrhythmias | 2208 (61.83) |
| 6 | Cardiovascular Disease (CVD) | 916 (25.65) |
| 7 | Chronic Obstructive Pulmonary Disease (COPD) | 1083 (30.33) |
| 8 | Coagulopathy | 593 (16.61) |
| 9 | Congestive Heart Failure (CHF) | 901 (25.23) |
| 10 | Deficiency Anemia | 681 (19.07) |
| 11 | Dementia | 195 (5.46) |
| 12 | Depression | 788 (22.07) |
| 13 | Diabetes | 1147 (32.12) |
| 14 | Drug Abuse | 311 (8.71) |
| 15 | Electrolyte Disorders | 1472 (41.22) |
| 16 | First Oxygen Levels, mean±s.d. | 96.27±5.35 |
|  | Gender |  |
| 17 | Male | 1907 (53.40) |
|  | Female | 1664 (46.60) |
| 18 | Hemiplegia | 162 (4.54) |
| 19 | Hispanic ethnicity | 944 (26.44) |
| 20 | Hypertension | 1965 (55.03) |
| 21 | Hypothyroidism | 510 (14.28) |
| 22 | Liver Disease | 778 (21.79) |
| 23 | Lymphoma | 83 (2.32) |
| 24 | Malignancy | 609 (17.05) |
| 25 | Metastatic Cancer | 204 (5.71) |
| 26 | Metastatic Solid Tumor | 204 (5.71) |
| 27 | Myocardial Infraction | 649 (18.17) |
| 28 | Obesity | 722 (20.22) |
| 29 | Other Neuro Disorders | 433 (12.13) |
| 30 | Paralysis | 165 (4.62) |
| 31 | Peptic Ulcers | 224 (6.27) |
| 32 | Peripheral Vascular Disease (PVD) | 729 (20.41) |
| 33 | Psychoses | 148 (4.14) |
| 34 | Pulmonary Circulation Disorders | 564 (15.79) |
| 35 | Race |  |
|  | White | 1605 (44.95) |
|  | African American | 1269 (35.54) |
|  | Other | 697 (19.52) |
| 36 | Renal Disease | 799 (22.37) |
| 37 | Renal Failure | 803 (22.49) |
| 38 | Rheumatic Disease | 171 (4.79) |
| 39 | Rheumatoid Arthritis | 232 (6.50) |
| 40 | Solid Tumor without Metastasis | 494 (13.83) |
| 41 | Valvular Disease | 758 (21.23) |
| 42 | Weight Loss | 574 (16.07) |
| *1 | Acute Respiratory Distress Syndrome (ARDS) | 260 (7.28) |
| *2 | COVID-19 Infection (+) | 789 (22.09) |
| *3 | Died | 293 (8.20) |
| *4 | ICU Admission | 963 (26.97) |

\* Outcome assessed

**Table S. 2.**
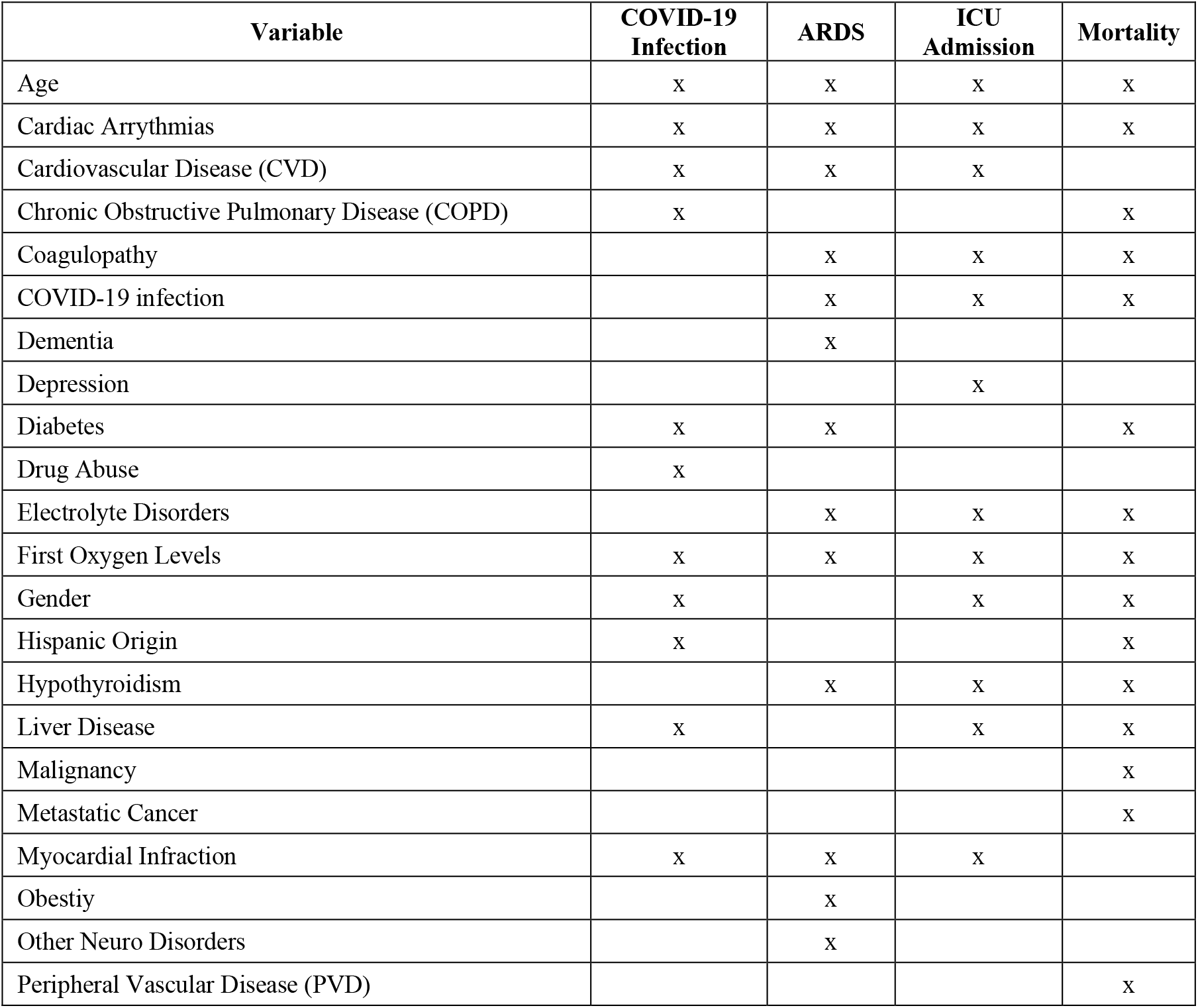
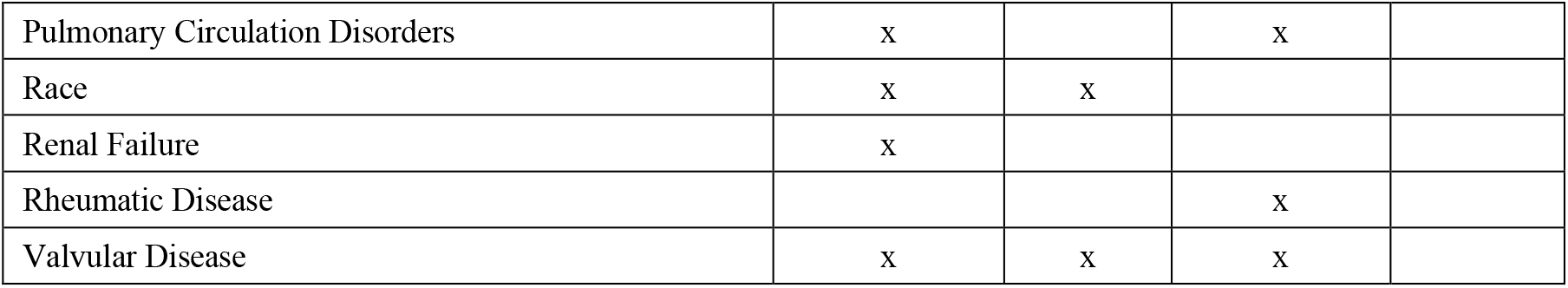
Clinical factors included in preliminary compact models (15 variables) to predict i) COVID-19, ARDS, ICU admission and risk of mortality.

**Figure S. 1.**
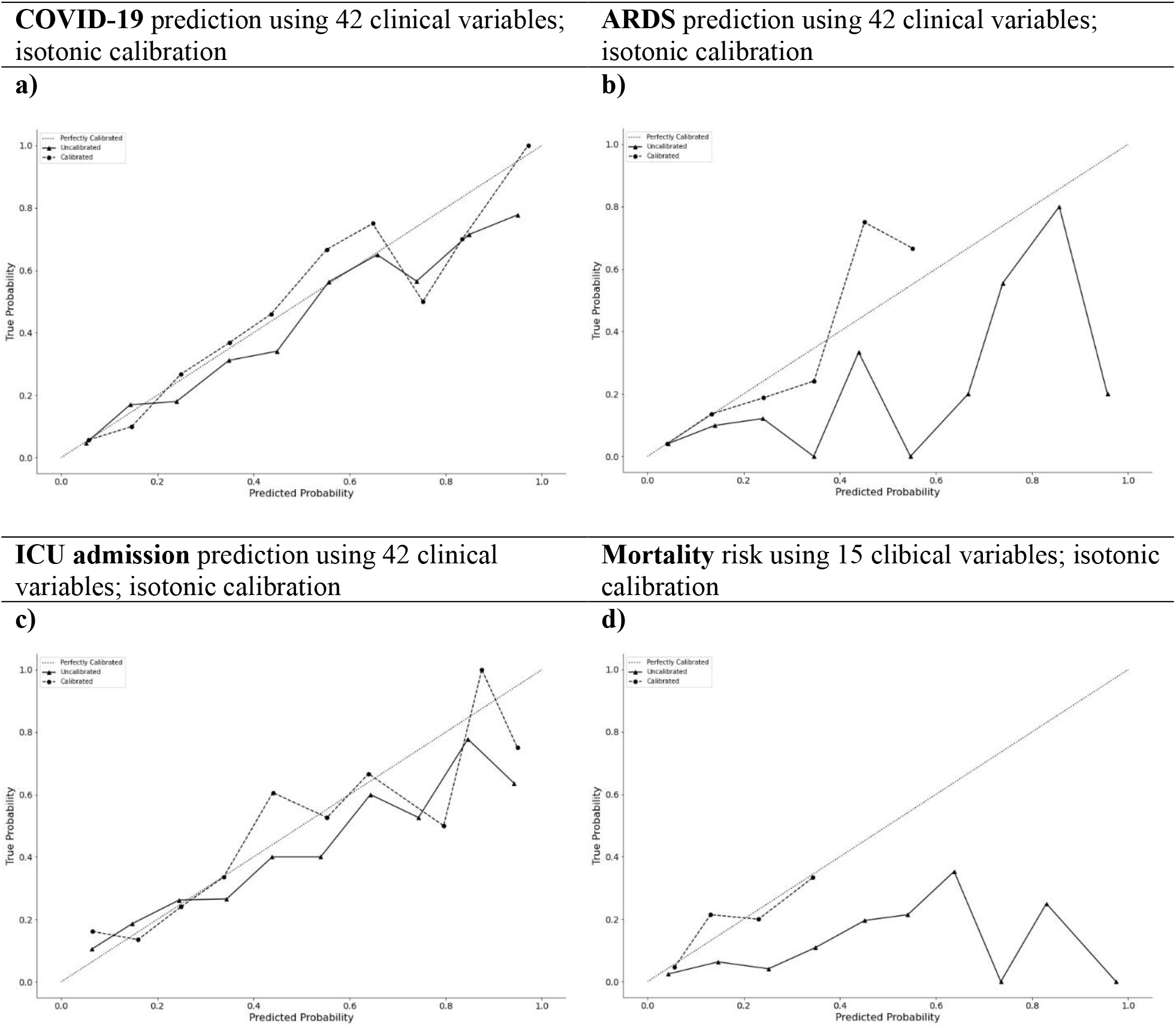
Calibration plots for uncalibrated and calibrated outcomes. Calibration compares true probability and predicted probabilities of a) COVID-19 infection, b) ARDS, c) ICU admission and d) risk of mortality (mortality). Uncalibrated classifiers are represented by a dotted line and calibrated classifiers are represented by a solid line.

## References

Azzam, H. C., Khalsa, S. S., Urbani, R., Shah, C. V., Christie, J. D., Lanken, P. N., & Fuchs, B. D. (2009). Validation study of an automated electronic acute lung injury screening tool. Journal of the American Medical Informatics Association, 16(4), 503–508.

Baud, D., Qi, X., Nielsen-Saines, K., Musso, D., Pomar, L., & Favre, G. (2020). Real estimates of mortality following COVID-19 infection. The Lancet infectious diseases, 20(7), 773.

Bhatnagar, V., Poonia, R. C., Nagar, P., Kumar, S., Singh, V., Raja, L., & Dass, P. (2020). Descriptive analysis of COVID-19 patients in the context of India. Journal of Interdisciplinary Mathematics, 1–16.

Castiglioni, I., Ippolito, D., Interlenghi, M., Monti, C. B., Salvatore, C., Schiaffino, S., Polidori, A., Gandola, D., Messa, C., & Sardanelli, F. (2020). Artificial intelligence applied on chest X-ray can aid in the diagnosis of COVID-19 infection: a first experience from Lombardy, Italy. MedRxiv.

Chavez, S., Long, B., Koyfman, A., & Liang, S. Y. (2020). Coronavirus Disease (COVID-19): A primer for emergency physicians. The American journal of emergency medicine.

Cucinotta, D., & Vanelli, M. (2020). WHO declares COVID-19 a pandemic. Acta Bio Medica: Atenei Parmensis, 91(1), 157.

Darby, A. C., & Hiscox, J. A. (2021). Covid-19: variants and vaccination. In: British Medical Journal Publishing Group.

DeLong, E. R., DeLong, D. M., & Clarke-Pearson, D. L. (1988). Comparing the areas under two or more correlated receiver operating characteristic curves: a nonparametric approach. Biometrics, 44(3), 837–845.

Dreher, M., Kersten, A., Bickenbach, J., Balfanz, P., Hartmann, B., Cornelissen, C., Daher, A., Stöhr, R., Kleines, M., & Lemmen, S. W. (2020). The characteristics of 50 hospitalized COVID-19 patients with and without ARDS. Deutsches Ärzteblatt International, 117(10), 271.

Gibson, P. G., Qin, L., & Puah, S. H. (2020). COVID-19 acute respiratory distress syndrome (ARDS): clinical features and differences from typical pre-COVID-19 ARDS. Med J Aust, 213(2), 54–56.

Gozes, O., Frid-Adar, M., Greenspan, H., Browning, P. D., Zhang, H., Ji, W., Bernheim, A., & Siegel, E. (2020). Rapid ai development cycle for the coronavirus (covid-19) pandemic: Initial results for automated detection & patient monitoring using deep learning ct image analysis. arXiv preprint 2003.05037.

Koenig, H. C., Finkel, B. B., Khalsa, S. S., Lanken, P. N., Prasad, M., Urbani, R., & Fuchs, B. D. (2011). Performance of an automated electronic acute lung injury screening system in intensive care unit patients. Critical care medicine, 39(1), 98–104.

Li, T., Han, Z., Wei, B., Zheng, Y., Hong, Y., & Cong, J. (2020). Robust screening of covid-19 from chest x-ray via discriminative cost-sensitive learning. arXiv preprint 2004.12592.

Ma, X., & Vervoort, D. (2020). Critical care capacity during the COVID-19 pandemic: global availability of intensive care beds. Journal of critical care, 58, 96.

Molnar, C. (2020). Interpretable machine learning. Lulu. com.

Niu, S., Tian, S., Lou, J., Kang, X., Zhang, L., Lian, H., & Zhang, J. (2020). Clinical characteristics of older patients infected with COVID-19: A descriptive study. Archives of gerontology and geriatrics, 89, 104058.

Rajpurkar, P., Irvin, J., Zhu, K., Yang, B., Mehta, H., Duan, T., Ding, D., Bagul, A., Langlotz, C., Shpanskaya, K., Matthew, & Andrew. (2017). CheXNet: Radiologist-Level Pneumonia Detection on Chest X-Rays with Deep Learning. arXiv pre-print server. https://doi.org/None 1711.05225

Ranieri, V. M., Rubenfeld, G. D., Thompson, B. T., Ferguson, N. S., Caldwell, E., Fan, E., Camprota, L., & Slutsky, A. S. (2012). Acute Respiratory Distress Syndrome: The Berlin Definition. Jama, 307(23), 2526–2533. https://doi.org/10.1001/jama.2012.5669

Robert, R., Coudroy, R., Ragot, S., Lesieur, O., Runge, I., Souday, V., Desachy, A., Gouello, J.-P., Hira, M., & Hamrouni, M. (2015). Influence of ICU-bed availability on ICU admission decisions. Annals of intensive care, 5(1), 1–7.

Wang, X., Peng, Y., Lu, L., Lu, Z., Bagheri, M., & Summers, R. M. (2017, 2017). ChestX-Ray8: Hospital-Scale Chest X-Ray Database and Benchmarks on Weakly-Supervised Classification and Localization of Common Thorax Diseases.

Weinacker, A. B., & Vaszar, L. T. (2001). Acute respiratory distress syndrome: physiology and new management strategies. Annual review of medicine, 52(1), 221–237.

WHO. (2021). WHO Coronavirus (COVID-19) Dashboard. Retrieved 13/04 from https://covid19.who.int/

Yadav, S., & Shukla, S. (2016). Analysis of k-fold cross-validation over hold-out validation on colossal datasets for quality classification. 2016 IEEE 6th International conference on advanced computing (IACC).

